# A Yield and Cost Comparison of TB Contact Investigation and Intensified Case Finding in Uganda

**DOI:** 10.1101/2020.03.12.20035071

**Authors:** Michael Kakinda, Joseph K. B Matovu

## Abstract

**Introduction:** Resource constraints in LMICs limit TB contact investigation despite evidence its benefits outweigh costs, with increased efficiency when compared with intensified case finding (ICF). However, there is limited data on the yield and cost per TB case identified in low resource-settings. We compared the yield and cost per TB case identified for ICF and TB-CI in Uganda.

**Methods:** A retrospective cohort study based on data from 12 Ugandan hospitals was done between April and September 2017. Two methods of TB case finding (i.e. ICF and TB-CI) were used. Regarding ICF, patients either self-reported their signs and symptoms or were prompted by health care workers, and those suspected to have TB were requested to produce a sputum sample. On the other hand, TB-CI was done by home-visiting and screening contacts of TB patients for TB; with those found with signs and symptoms requested to produce sputum samples for examination. TB yield was defined as the ratio of diagnoses to tests, and this was computed per method of diagnosis. The costs per TB case identified (medical, personnel, transportation and training) for each diagnosis method were computed using the activity-based approach, from the health care perspective. Cost data were analyzed using Windows Excel.

**Results:** 454 index clients’ cases and 2,707 of their household contacts were investigated. Thirty-one per cent of contacts (840/2707) were found to be presumptive TB cases. A total of 7,685 tests were done, 6,967 for ICF and 718 for TB-CI. ICF had a yield of 18.62% (1297/6967) at a cost of USD $120.60 to diagnose a case of TB while TB-CI had a yield of 5.29% (38/718) at an average cost of USD $ 877.57 to diagnose a case of TB.

**Conclusion:** Regarding case-finding, the yield of TB-CI was four-times lower and seven-times costlier compared to ICF. These findings suggest that ICF can improve TB case detection at a low cost, particularly in high TB prevalent settings.

## BACKGROUND

Household contacts of pulmonary bacteriologically-confirmed (PBC) tuberculosis (TB) patients have a high risk of becoming infected with *Mycobacterium tuberculosis* and developing active TB (1,2,3,4). Investigation of these contacts for TB could lead to early detection and treatment of active TB as well as prophylactic treatment of persons with latent TB (5,6). This has long been a priority intervention in most low-incidence countries to reduce TB incidence (7). However, in low and middle-income countries (LMICs), home visits for TB contact investigation (TB-CI) are limited by the high workload of health workers, poor adherence to treatments for latent TB and resources (2,8). But the stagnating Case Detection Rates (CDR) in dual epidemic TB and HIV countries (9,10) and increasing drug-resistant TB have prompted a reassessment of the potential benefit of TB-CI (11).

A recent TB-CI review conducted in LMICs found a cumulative yield of 4.5% (95% CI 4.3%-4.8%) among household contacts (11), suggesting’s that it has a utility in detecting additional cases. Mathematical models have suggested that the benefits of TB-CI could out-weigh costs and even increase efficiency in the long-run when compared with passive case finding (PCF) (12,13). With TB-CI, TB cases are likely to be detected early, thereby minimizing the likelihood of further complications and hospitalization (14). It is also possible to assume that with early TB case detection, TB transmission is interrupted and further cases are prevented (15). TB-CI also removes some barriers to health care since health workers reach out to potential TB patients in the community, removing the pre-treatment costs and increasing access to health services (16).

Furthermore, empirical evidence of the cost-effectiveness of implementing TB-CI in real-world settings is limited (8,17-21). To date, only Ssekandi et al (8) have evaluated the cost and yield of various TB case finding strategies in Africa. This study therefore compared TB yield and cost for ICF and TB-CI in an operational setting in Uganda-a high burden TB/HIV country in sub-Saharan Africa.

## METHODS

### Study Design and Setting

This was a retrospective cohort study of TB-CI using data collected from 11 Regional Referral Hospitals and one district Hospital in Uganda (Table 1). By the Uganda Ministry of Health categorization, Regional Referral Hospitals (RRHs) serve a population of 3 million persons while General Hospitals (GHs) serve a population of 500,000 people (22).

**TABLE 1:** Description of the Study Sites

| Health Facility | Bed Capacity | Annual OPD<sup>#</sup> Attendance (2017) | Distance & Direction from Capital |
| --- | --- | --- | --- |
| Arua RRH | 323 | 153,451 | 480 km NW |
| Gulu RRH | 397 | 97,415 | 333.7 km N |
| Jinja RRH | 408 | 165,573 | 87 km E |
| Hoima RRH | 268 | 41,930 | 190 km W |
| Kabale RRH | 226 | 63,266 | 426 km SW |
| Kawolo GH | 106 | 51,946 | 48.5 km E |
| Lira RRH | 346 | 95,695 | 340 km N |
| Masaka RRH | 540 | 125,025 | 130.2 km SSW |
| Mbale RRH | 355 | 56,539 | 244 km E |
| Moroto RRH | 172 | 54,188 | 463 km NE |
| Mubende RRH | 173 | 45,681 | 149.1 km WSW |
| Soroti RRH | 251 | 90,818 | 298 km NNE |

\*RRH-Regional Referral Hospital, \*GH-General Hospital, #OPD-Out Patient Department

### Study Population

The study population was pulmonary bacteriologically-confirmed (P-BC) tuberculosis patients (aged 18+ years) that were diagnosed at the above-mentioned study sites between April and September 2017. All consented PBC TB patients were visited at home for TB-CI. A household contact was defined as an individual who had resided in the household of an index TB patient for at least 7 consecutive days during the 3 months prior to the diagnosis of TB in the index case.

### TB Diagnosis

TB case finding was conducted using two different methods; Intensified Case Finding (ICF) and TB contact investigation (TB-CI). These methods are described in detail below.

#### a) Intensified Case Finding (ICF)

Under ICF, individuals with TB symptoms who self-report to health facilities are assessed for TB using the Intensified Case Finding Tool. It assesses for a cough lasting 2 weeks or more or any a cough for people living with HIV (PLHIV) or any of these signs and symptoms (evening persistent fevers > 2 weeks, excessive night sweats > 3 weeks and noticeable weight loss) were presumed to have TB and were subjected to further investigations to ensure that a definitive diagnosis was done. TB was diagnosed on receiving a *Mycobacterium Tuberculosis* detected test from the GeneXpert test.

#### b) TB Contact Investigation (TB-CI)

TB patients that consented to a home visit were visited by two health workers, a nurse and a community health worker. Upon reaching the home, the team offered health education to household members.After family members were screened for TB using the Intensified Case Finding (ICF) Tool as above. Any household contact who reported any of the signs and nsymptoms in the ICF tool was categorized as a presumptive TB case. Individuals who were found to be coughing were requested to provide a sputum sample which was transported to the health facility and subjected to MTB RIF GeneXpert (Cepheid, Sunnyvale, CA, USA). The other presumptive TB cases were referred to the health facility for further clinical examinations.

## Data Collection Methods and Procedures

### Intensified Case Finding Data

Intensified case finding (ICF) data, included the total number of cases, number of PBC cases, number of MTB/RIF GeneXpert tests done to detect those cases. These data were extracted from the program registers and placed in a database.

### TB Contact Investigation (TB-CI) Data

TB-CI data were extracted using a data extraction tool from health facility program data. Variables extracted from the records included patient’s TB numbers, age, sex, number of contacts, number of contacts presumed to have TB, number of contacts offered a definitive diagnostic test for TB, number of contacts with TB, number of contacts who had an HIV test, and number of patients found HIV positive. These data were then entered into a spreadsheet (Windows Excel 2013, Microsoft Corp., Redmond, WA) in preparation for analysis.

### Costing Data

Costing data were collected on program costs including personnel, transport and training costs, as described below. We did estimate the direct economic costs related to diagnosing a TB case using either ICF or TB-CI from a provider’s perspective. Most of the information was extracted from program records, together with interviews with staff at the various health care facilities. Efforts were made to adhere to guidelines stipulated in literature (23,24). We were not able to estimate the overhead costs such as utilities, custodial services, buildings, office space, computers and maintenance of medical equipment. These costs were excluded because we could not tease them out of the costs associated with the provision of other health care services (25).

#### a) Personnel Costs

Personnel costs were estimated per method of TB diagnosis, as described below.

##### i) Intensified Case Finding (ICF)

We collected data on personnel time spent by Nurses, Nursing Assistants, Clinicians (Clinical Officers and Medical Officers) and Laboratory Technologists. Using a time-series, ten samples were done per carder per health facility and the average taken.

The hourly rates were calculated from monthly salaries as paid by the government of Uganda in 2017 assuming a 40 hours working week (26). The total personnel costs were obtained by multiplying the hourly rate with the estimated patient contact time, the assumption was every presumptive TB case made it to the laboratory. Hence the number of tests done was used as a proxy for patients seen by the health care workers for TB case finding.

##### ii) TB Contact Investigation (TB-CI)

A stop-clock was used to obtain the time it took 2 health care workers to travel from the health facility to the patient’s home, conduct contact investigation and back. To calculate the personnel cost for TB-CI the same method as above was used, factoring in the hourly pay and time for the activity. The other costs incurred were meals and incidental expenses for the persons who performed TB-CI. USD $ 5.56 was given per day, per person when they did perform TB-CI activities as per the guidance (27).

#### b) Transportation Costs

Transportation costs were incurred when the Health Care Workers went to perform contact investigation. The distance between the Health Facility and Index patient’s home was calculated from the odometer of a motorbike or a motor vehicle which was used for transport. The health care worker and a community health worker were compensated with UGX 200 (USD 0.056) per kilometer as is the guidance (27).

#### c) Training Costs

24 health care workers were trained, 2 per health facility, a clinician and a nurse for 5 days. The costs incurred were their per diems and transport. The per diem was USD $ 44.78 per participant and facilitator as directed by standing orders for the government of Uganda per night (27) Transport allowance was by public means for the participant and was calculated depending on the distance from the health facility where the participant is based on the training venue. The participant was compensated for UGX 200 (USD 0.056) per kilometer. Other costs incurred included the cost of hiring the training venue, payment for meals and refreshments during training and fuel for facilitators. These were all computed to get the total training costs. To compute the unit cost for training per TB case diagnosed by contact investigation, we divided the total costs of the training by TB cases diagnosed by contact investigation. There were no training costs incurred for ICF.

#### d) Medical Costs

Medical costs were defined as all costs incurred at the point where health care was delivered. This included GeneXpert MTB/RIF and all other consumables required. The unit cost of an MTB/RIF GeneXpert test per patient was obtained from the literature. A GeneXpert test was found to cost on average 21 USD in Uganda (28). The number of tests done under each approach was obtained from program data, and these were 6967 tests for ICF and 718 for TB-CI. The costs of consumables were market-based and were obtained from the National Medicines Stores (NMS) catalogue (29). NMS is a government of Uganda agency responsible for the supply of medicines and other related products throughout the country. The quantities of consumables were calculated based on what a health care worker would use for a single patient.

### Study Outcomes

The primary outcome was TB yield in contacts of PBC TB patients contacted at home compared with those who self-referred themselves to the health facilities. The secondary outcome was the cost per TB case diagnosed from TB-CI when compared with the cost per TB case diagnosed using ICF.

### Data Analysis

We exported data from a spreadsheet (Microsoft Excel version 2013) into Intercooled Stata version 13 (Stata-Corp, College Station, Texas, USA). We summarized demographic and outcome data into frequencies, percentages, and measures of central tendency, the median and mean. The yield of TB was obtained by dividing the number of TB cases from contact investigation divided by the number of tests done.

The cost per TB case identified by TB-CI was calculated from the healthcare perspective. All the ingredients needed to diagnose a TB case, using either ICF or TB-CI, were considered. These were summed up to get the total cost of diagnosing a TB case by either ICF or TB-CI.

To determine the unit cost of either diagnosing a TB case using either ICF or TB-CI, the total costs of either modality were divided by the TB cases obtained from each method either ICF or TB-CI. The costs were converted from Uganda Shillings to 2017 United States Dollars (USD) rate at an exchange rate of 1 USD to 3595 Uganda Shillings, there was no discounting since the duration was less than a year.

### Ethical Approval

ICF is part of routine clinical care, while TB-CI is a WHO/Ministry of Health in Uganda recommended strategy and does not require ethical approval for introduction into a program. The secondary analysis was done on aggregate and anonymous project data, therefore there was no need for additional ethical clearance. But consent was sort from the patients prior to the home visit for contact investigation.

## RESULTS

### Participants’ Characteristics

The 454 index TB patients had a total of 2,707 contacts, with an average household size of six members (SD: ±8.70). The mean age of index patients and contacts were not very different with 33.6 years and 34.7 years respectively. The index TB patients were most likely to be males (65.2%) while there was an equal distribution of sex among the contacts with males being 50.97%. 37.4% (170/454) of the index Patients had HIV while only 3.97 % (74/1864) of the contacts had HIV (See Table 2).

**TABLE 2:** Socio-demographic Characteristics of Index Patients and their Household Contacts

| INDEX TB PATIENTS |  |  |
| --- | --- | --- |
| Number of Index TB Patients | Total (N) | 454 |
| Age | Average | 33.6 (SD = 13.734) |
|  | Median | 32 (0-90) |
| Sex | Female | 158 (34.80%) |
|  | Male | 296 (65.19%) |
| HIV Status | HIV-positive | 170 (37.44%) |
|  | HIV-negative | 284 (62.55%) |
| HOUSEHOLD CONTACTS |  |  |
| Number of Contacts | Total (N) | 2707 |
|  | Average | 6.0 (SD = 8.703) |
|  | Median | 4.0 (0-125) |
| Age | Average | 34.7 (SD-12.45) |
|  | Median | 31 (0-84) |
| Sex | Female | 1327 (49.03%) |
|  | Male | 1380 (50.97%) |
| HIV status | HIV-positive | 74 (2.73%) |
|  | HIV-negative | 1790 (66.12%) |
|  | Unknown | 843 (31.14%) |

**Table 3** shows the number of patients notified, the cases investigated at home and TB yield at each study site. 6967 MTB RIF GeneXpert tests were done to diagnose 1297 TB cases resulting in an ICF yield of 19% (1297/6967). Of the 1,295 Pulmonary Bacteriologically-Confirmed (PBC) TB cases, 454 (35.0%) had their contacts investigated for TB.

**TABLE 3:** Number of patients notified, cases traced at home and TB yield at each study site

| HEALTH FACILITY | NOTIFIED TB CASES (APRIL-SEPT'17) | MTB/RIF GeneXpert Tests-ICF | P-BCβ CASES (APRIL-SEPT'17) | TB Yield for ICF | INDEX Pts $\gamma$ | NUMBER OF HOUSEHOLD CONTACTS | PRESUMED TB | INVESTIGATED FOR TB | TB Pts $\gamma$ | TB Yield for HCI |
| --- | --- | --- | --- | --- | --- | --- | --- | --- | --- | --- |
| Arua RRH* | 353 | 561 | 168 | 30% | 51 | 629 | 210 | 190 | 15 | 7.89% |
| Gulu RRH* | 221 | 660 | 131 | 20% | 24 | 140 | 12 | 12 | 1 | 8.33% |
| Hoima RRH* | 273 | 1253 | 111 | 9% | 26 | 69 | 12 | 12 | 0 | 0.00% |
| Jinja RRH* | 342 | 404 | 84 | 21% | 30 | 58 | 17 | 17 | 0 | 0.00% |
| Kabale RRH* | 86 | 697 | 66 | 9% | 20 | 69 | 4 | 4 | 1 | 25.00% |
| Kawolo GH# | 110 | 314 | 45 | 14% | 48 | 249 | 64 | 35 | 2 | 5.71% |
| Lira RRH* | 271 | 669 | 178 | 27% | 107 | 710 | 220 | 218 | 9 | 4.13% |
| Masaka RRH* | 297 | 527 | 139 | 26% | 18 | 72 | 5 | 4 | 0 | 0.00% |
| Mbale RRH* | 180 | 611 | 104 | 17% | 13 | 45 | 12 | 12 | 0 | 0.00% |
| Moroto RRH* | 200 | 454 | 109 | 24% | 23 | 114 | 38 | 37 | 4 | 10.81% |
| Mubende RRH* | 137 | 569 | 96 | 17% | 58 | 296 | 204 | 139 | 3 | 2.16% |
| Soroti RRH* | 121 | 248 | 66 | 27% | 36 | 256 | 42 | 38 | 3 | 7.89% |
|  | 2591 | 6967 | 1297 | 19% | 454 | 2,707 | 840 | 718 | 38 | 5.29% |

\*RRH-Regional Referral Hospital, # GH-General Hospital, <sup>†</sup>P-BC-Pulmonary Bacteriologically Confirmed, <sup>‡</sup> Pts-Patients

Only 31.0% (840/2,707) were presumed to have TB. 85.5% (718/840) of the presumptive TB contacts had their samples sent to the laboratory for a definitive diagnosis. Of these, 5.3% (38/718) (95% CI 3.8% −7.2%) were diagnosed with TB (Table 3). The Number Needed to Screen (NNS) to get one TB case was 71 (718/38). So, on average contact tracing, 100 index cases will lead to 8.4 (38/454*100) additional diagnoses.

Kabale regional referral hospital had the highest TB yield (25%) followed by Moroto regional referral hospital (10.8%) while Jinja, Hoima, Masaka and Mbale had the lowest TB yield with no patients diagnosed with TB during TB-CI.

### Cost for identifying a TB case using different case-finding modalities

Table 4 shows the cost of finding a TB Case using either ICF or TB-CI. The total costs were USD $ 156,416.62 and USD $ 33,347.83 for ICF and TB-CI respectively, while the unit costs were USD $ 120.60 for ICF and USD $ 877.57 for TB-CI. Under ICF, 96.18% of the cost went to performing MTB/RIF GeneXpert Tests while 2.64% was to Consumables and Supplies and 3.82% went to salaries. The main driver of costs under TB-CI was training at 30.71%; GeneXpert tests at 40.21%; allowances & transport at 20.21%; and supplies and consumables at 0.34%.

**Table 4:** Cost of finding a TB Case using either Passive Case Finding (ICF) or TB Contact Investigation

| COST CATEGORY | CASE FINDING MODALITY |  |  |  |
| --- | --- | --- | --- | --- |
|  | ICF* |  | TB-CI# |  |
| PROGRAM COSTS | Cost | % Cost | Cost | % Cost |
| Personnel Costs |  |  |  |  |
| Training | 0 | 0.00% | 10,242.16 | 30.71% |
| Salaries | 5,980.99 | 3.82% | 1,174.73 | 3.52% |
| Allowances & Transport Costs | 0 | 0.00% | 6,738.04 | 20.21% |
| Sub-Total-Personnel Costs | 5,980.99 | 3.82% | 18,154.93 | 54.44% |
| **Medical Costs** |  |  |  |  |
| Supplies & Consumables | 4,128.63 | 2.64% | 114.9 | 0.34% |
| MTB RIF GeneXpert | 146,307.00 | 93.54% | 15,078.00 | 45.21% |
| Sub-Total-Medical Costs | 150,435.63 | 96.18% | 15,192.90 | 45.56% |
| **TOTAL** | **156,416.62** | **100.00%** | **33,347.83** | **100.00%** |
| Total TB Cases (PBC) | 1,297.00 |  | 38 |  |
| **Cost per case** | **\$120.60** | | **\$877.57** | |

(\*ICF-Intensified Case Finding, #TB-CI-TB Contact Investigation)

## DISCUSSION

We found TB-CI had a yield four times lower than ICF (5.29% versus 18.61%), and it cost almost seven-times more (USD $ 877.57 versus USD $ 120.60) to diagnose a case of TB using TB-CI when compared to ICF, suggesting that use of ICF is a more cost-effective strategy in identifying new TB cases particularly in high TB prevalent settings such as Uganda.

Our TB-CI yield despite being four times lower than ICF, it is higher than the literature findings whose estimates are about half what we got especially for PBC cases (8, 30). Uganda is one of the 30 high TB/HIV burden countries, thus, the high TB yield observed across the country could be a clear testimony to the already high prevalence of TB in Uganda as reported from previous studies (31). It is important to note that there were variations reported by region with the highest TB yield reported in Kabale Regional Referral Hospital. Our study did not explore the reasons for the high TB yield in Kabale Regional Referral Hospital. However, this could be due to the late adoption of the intervention by some sites compared to others after training. Nevertheless, finding a TB yield of 5.29% for PBCs only does suggests that TB-CI is a worthwhile intervention that can be used to increase TB cases detection.

We found that it cost seven-times more to diagnose a case of TB using TB-CI compared to ICF. Our TB-CI costs were higher than those reported by Sekandi et al who found that it cost 416.35 USD (10). This is maybe due to the fact that Sekandi et al used Smear microscopy which costs on average USD $ 1-3, while we used MTB/RIF GeneXpert which costs on average USD $ 21 (range USD $ 16-58) (28). The other probably reason is our study was of a shorter duration (i.e. 6 months) compared to the duration of the study conducted by Sekandi et al which lasted 18 months. The longer study duration could have helped the study team to diagnose more TB cases which could have reduced the cost per case. Our study was also more extensive – i.e. it included data collected from 12 health facilities across the country yet the study by Ssekandi only covered the capital city, Kampala. Despite the higher cost, TB-Contact Investigation should be a worthwhile intervention given that patients are likely to be diagnosed early and hence decreasing their morbidity and mortality, and there are likely to be other benefits like a reduced stigma to the patient since the household has a better understanding of the disease. But, it may be helpful to integrate TB-CI with other interventions (such as immunization, malnutrition, family planning or HIV testing) to reduce the cost but also provide more holistic care to families and communities.

Our retrospective observational study was not without limitations. First and foremost, we did use some costs that were borrowed from the available literature and there could have been a publication bias since we did not collect the said data ourselves. It is also worth noting that the data were mainly collected for operational purposes and not for research. Therefore, some data were missing; we tried to circumvent this by corroborating information from a patient’s clinical notes and excluding the missing data from the analysis. Secondly, since this was in an operational setting, we did not have access to sputum cultures and chest radiographs for most of the presumptive TB Cases. Only patients that were pulmonary bacteriologically confirmed using the MTB RIF GeneXpert were considered. The TB cases diagnosed with TB contact investigation could have been more had other diagnostic modalities been considered.

## CONCLUSIONS

The yield of TB-CI was four-times lower than that of ICF, but it cost seven-times more to identify a single case of TB under TB-CI than ICF. These findings suggest that ICF can improve TB case detection at a low cost, particularly in high TB prevalent settings.

## Data Availability

All the data is available, but it is mainly costing data

## Notes

### Competing Interest Statement

The authors have declared no competing interest.

### Clinical Trial

The study did not require ethical approval, it was part of routine care and anonymous data was used

### Funding Statement

There was no funding obtained.

